# Misidentification of *Plasmodium* mixed-infections leads to an overestimation of falciparum malaria

**DOI:** 10.1101/2023.10.02.23296210

**Authors:** Nimita Deora, Veena Pande, Abhinav Sinha

## Abstract

Despite the fact that malaria elimination is nearing in several countries, we continue to struggle with accurate diagnosis and thus treatment. The purpose of this study was to determine the rate of *Plasmodium* species misidentification (MI) by microscopy (MS). The study was based on previously published reports in which MS-PCR pairs were analysed to identify *Plasmodium* misidentification rates (by MS). Region- and species-wise misidentification rates were also estimated.

A total of 2706 MS-PCR pairs were extracted from 16 different locations across 11 Indian states. MS-PCR pair analysis revealed 15% misidentification rate (408/2706). Surprisingly, microscopy misidentified more than 98% of mixed infections (400/405) as mono-infections (almost all as *P. falciparum* mono infections). The study identifies Jharkhand and Madhya Pradesh as major contributors (>20%) to *Plasmodium* species misidentification by microscopy. These findings suggest that we are overestimating *P. falciparum* burden, potentially wasting elimination resources, and underestimating non-falciparum species. The study also addresses an important issue concerning analysis of misidentification & sub-microscopic infection data (SMI). The proposed analysis (for MI and SMI) will aid in deciphering MI and SMI data in a more granular manner, generating actionable data for elimination programmes in various countries.

## Introduction

Malaria is a disease caused by species of genus *Plasmodium* which is transmitted by female Anopheles mosquitoes. Globally, 247 million people over 84 countries were estimated to have malaria with 619 000 estimated deaths in 2021. The WHO South East Asia Region had only ∼2% of the estimated global burden, but India contributed to >75% of these cases with *P. vivax* and *P. falciparum* as major species with negligible reporting of the non-Pv and non-Pf parasite species.^1^ Therefore, the current recommended antimalarial in India are targeted for Pf (ACTs; AS+SP and AL) and Pv (chloroquine). Additionally, radical 14-day treatment with primaquine is recommended for Pv. All mixed infections involving Pf are recommended to be treated as Pf with additional 14-day treatment with primaquine if the other species is Pv and/or Po. For other mixed infections, ACTs should be used.^2^ The use of primaquine is recommended only after ensuring G6PD sufficiency. For malaria diagnosis, the country relies on microscopy (malaria +/- following examination of stained thick and thin peripheral blood smears from febrile persons) and bivalent RDTs.

Microscopy (MS) still remains the ‘gold standard’ for malaria diagnosis and surveillance in field settings owing to it being relatively cheaper and technologically simpler. Under normal field conditions, the limit of detection (LoD) of MS hovers around ≥100 parasites/μl of blood, whereas a skilled microscopist can detect as few as 5-10 parasites/μl. In contrast, PCR-based methods have LoD between 0.022 (ultrasensitive PCR) and 5 (standard PCR) parasites/μl of blood.^3–5^

It is true that some *Plasmodium* species, are visually so similar that it is difficult to discriminate Po from Pv^6–7^ and Pm from Pk^6–8^ using MS thus underestimating the real species-specific burden of *Plasmodium*. MS is a skill-based technique and it necessitates quality assurance from smear preparation to slide examination.^9–13^ As lab technicians have difficulty in diagnosing Pm, Po, and Pv, non-falciparum species may be misdiagnosed, especially when present with Pf during mixed infections.^14^ In such cases, treatment is provided for Pf alone (most likely ACT), whereas co-infecting species like Pv and Po remain untreated which should be additionally treated with primaquine to avoid relapses. Therefore, for a particular species, false-negative microscopy (misidentified as another species) could result in potentially serious consequences. Accurately distinguishing between Pf and non-Pf infections and identifying all *Plasmodium* species (present in a region), is a minimum competence required for basic MS.^13^

Since MS is still the diagnostic “gold standard”, its sensitivity and specificity are vital parameters for countries aiming malaria elimination. Compromised sensitivity (false negative/FN or sub-microscopic infections/SMIs) and specificity (false positive/FP or misidentified *Plasmodium* species/MI) may be detrimental. Despite the gravity, targeted research on these issues has rarely been a priority and therefore the way FN/SMI data are analyzed and reported tends to be flawed. Deep understanding and analysis of microscopy and PCR paired data is therefore critical as most of the published reports interpret this data from flawed and crude analysis which may lead to incorrect conclusions.

In order to address the above concerns the current study attempts to depict the magnitude of *Plasmodium* species misidentification in India and whether misidentification of mixed infections is associated only with uncommon/rare *Plasmodium* species. This manuscript also cautions and critiques ‘overall’ analysis and proposes a one-to-one analysis of microscopy-PCR paired data which is more factual and reveals much needed information for malaria control, globally. The suggested analysis is expected to reveal more granular and actionable information that would be immensely helpful for the National authorities driving malaria elimination in various countries.

## Methodology

Studies that were included for this study were selected from a database that had previously been created to assess the worldwide burden of mixed *Plasmodium* species infection. The study’s protocol was registered into the International Prospective Register of Systematic Reviews (PROSPERO) with the registration number CRD42021234278.

From the above database, studies that contained data from India were included if MS and PCR both were applied on the same set of samples for diagnosing *Plasmodium spp* (Figure 1).

**Figure 1:**
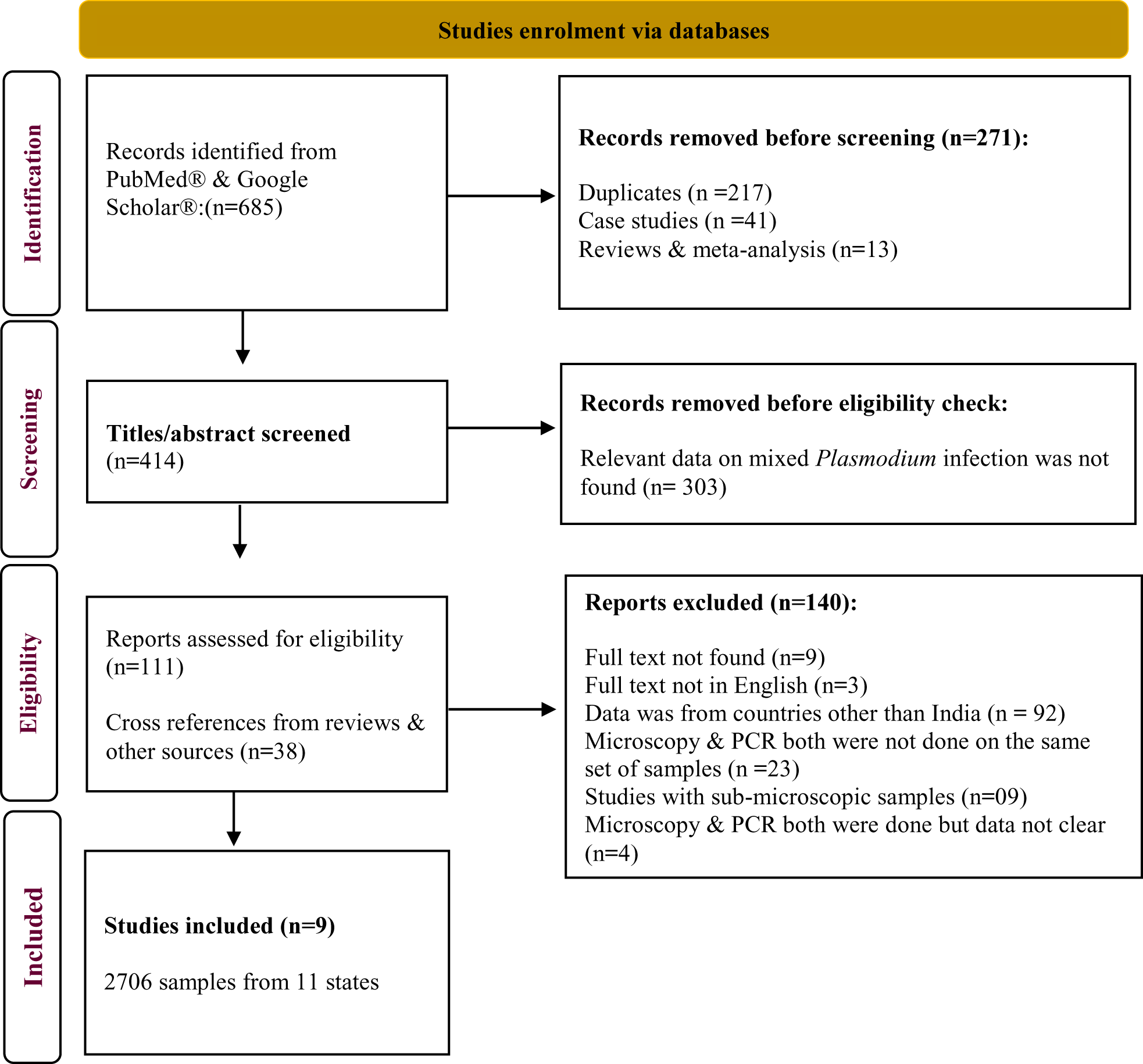
Study enrolment following PRISMA guidelines.

From the enrolled studies, the year of data-collection, geographical area, total number of samples tested, number of samples positive/negative for the presence of *Plasmodium spp.* by MS and PCR (concordant and discordant results) were recorded (Table 1). Samples were analysed by data collection sites and if the same data collection sites were found in numerous studies, they were not grouped together but were examined individually. Misidentified (MI) *Plasmodium* species (misdiagnosed by MS as a different *Plasmodium* species) were identified. Samples that were detected positive by MS but negative by PCR for all *Plasmodium* species were also categorized as misidentified.

**Table 1:** The table displays misidentification (species-wise) data (number of cases detected by MS and PCR) from 11 states derived from extracted reports, with the top row indicating the year of data collection. In the second row, states were arranged alphabetically & then according to year of data-collection in ascending order. MS: Microscopy, PCR: Polymerase Chain Reaction, AP: Arunachal Pradesh, AS: Assam, CH: Chhattisgarh, GUJ: Gujarat, JH: Jharkhand, KA: Karnataka, MP: Madhya Pradesh, MH: Maharashtra, OD: Odisha, RJ: Rajasthan, TR: Tripura

| Year → | 2005 |  | 2011 |  | 2010 |  | 2013-14 |  | 2014 |  | 2015 |  | 2014 |  | 2014 |  | 2012-14 |  | 2012-15 |  | 2012 |  | 2014 |  | 2014 |  | 2014 |  | 2014 |  | 2014 |  |
| --- | --- | --- | --- | --- | --- | --- | --- | --- | --- | --- | --- | --- | --- | --- | --- | --- | --- | --- | --- | --- | --- | --- | --- | --- | --- | --- | --- | --- | --- | --- | --- | --- |
| State → | AP |  | AS |  | CG |  | CG |  | CG |  | CG |  | GUJ |  | JH |  | KA |  | KA |  | MP |  | MP |  | MH |  | OD |  | RJ |  | TR |  |
| Diagnosis → | MS | PCR | MS | PCR | MS | PCR | MS | PCR | MS | PCR | MS | PCR | MS | PCR | MS | PCR | MS | PCR | MS | PCR | MS | PCR | MS | PCR | MS | PCR | MS | PCR | MS | PCR | MS | PCR |
| Pf |  |  | 53 | 48 | 256 | 253 | 214 | 188 | 200 | 199 | 355 | 284 | 97 | 87 | 216 | 158 |  |  |  |  |  |  | 226 | 174 | 234 | 211 | 267 | 219 | 140 | 112 | 127 | 107 |
| Pv |  |  |  |  |  |  |  |  |  |  |  |  |  |  |  |  | 124 | 116 | 161 | 116 |  |  |  |  |  |  |  |  |  |  |  |  |
| Pm | 8 | 7 | 0 | 1 |  |  |  |  |  |  |  |  |  |  |  |  |  |  |  |  | 22 | 14 |  |  |  |  |  |  |  |  |  |  |
| PfPv |  |  | 5 | 4 |  |  | 0 | 23 |  |  | 0 | 59 | 0 | 8 | 0 | 55 | 0 | 8 | 0 | 45 |  |  | 0 | 47 | 0 | 21 | 0 | 40 | 0 | 26 | 0 | 19 |
| PfPm | 1 | 1 | 0 | 3 |  |  | 0 | 2 |  |  | 0 | 3 | 0 | 2 |  |  |  |  |  |  | 0 | 3 | 0 | 4 | 0 | 2 | 0 | 6 | 0 | 2 | 0 | 1 |
| PfPo |  |  |  |  | 0 | 2 | 0 | 1 |  |  |  |  |  |  | 0 | 2 |  |  |  |  |  |  | 0 | 1 |  |  | 0 | 2 |  |  |  |  |
| PvPm | 0 | 1 |  |  |  |  |  |  |  |  |  |  |  |  |  |  |  |  |  |  | 0 | 2 |  |  |  |  |  |  |  |  |  |  |
| PfPmPo |  |  |  |  |  |  |  |  |  |  |  |  |  |  | 0 | 1 |  |  |  |  |  |  |  |  |  |  |  |  |  |  |  |  |
| PfPvPm |  |  |  |  |  |  |  |  |  |  | 0 | 5 |  |  |  |  |  |  |  |  |  |  |  |  |  |  |  |  |  |  |  |  |
| PfPvPo |  |  |  |  | 0 | 1 |  |  | 0 | 1 | 0 | 1 |  |  |  |  |  |  |  |  |  |  |  |  |  |  |  |  |  |  |  |  |
| PfPvPmPo |  |  |  |  |  |  |  |  |  |  | 0 | 1 |  |  |  |  |  |  |  |  |  |  |  |  |  |  |  |  |  |  |  |  |
| Negative by MS (for all spp.) |  |  | 0 | 2 |  |  |  |  |  |  | 0 | 2 |  |  |  |  |  |  |  |  | 0 | 3 |  |  |  |  |  |  |  |  |  |  |
| Total | 9 | 9 | 58 | 58 | 256 | 256 | 214 | 214 | 200 | 200 | 355 | 355 | 97 | 97 | 216 | 216 | 124 | 124 | 161 | 161 | 22 | 22 | 226 | 226 | 234 | 234 | 267 | 267 | 140 | 140 | 127 | 127 |

All mixed *Plasmodium* species infections that were misidentified as mono-infection (by MS) of either species were also categorized under MI as it was uncertain if the other species (in a mixed-infection) was missed by MS because it was sub-microscopic or because the other one was detected sooner.

Burden of overall and species-specific *Plasmodium* MI was estimated as:

A. **Overall & region-specific MI rate (%)** = total MI cases by MS (irrespective of species) / total cases identified by MS) * 100
B. **Species-specific MI rate (%)** was estimated as:

**(i)** total MI cases by MS (of a particular species) / total cases identified by MS) * 100
**(ii)** total MI cases by MS (of a particular species) / total MI cases (irrespective of species) * 100
**(iii)** total MI cases by MS (of a particular species) / total PCR confirmed cases (of that particular species) * 100

## Results

The study finally included 9 published records^24^ (Figure 1) encompassing 2706 MS-positive *Plasmodium* samples collected between 2005 and 2015 from 11 states and 16 sites (Table 2). Overall MI rate was 15% (408/2706) including negative samples (n=7; MS-positive but PCR-negative for all *Plasmodium* species). Further analyses revealed that all (99.75%; 400/401) MI samples were mixed-infections except one mono-infection.

**Table 2:**
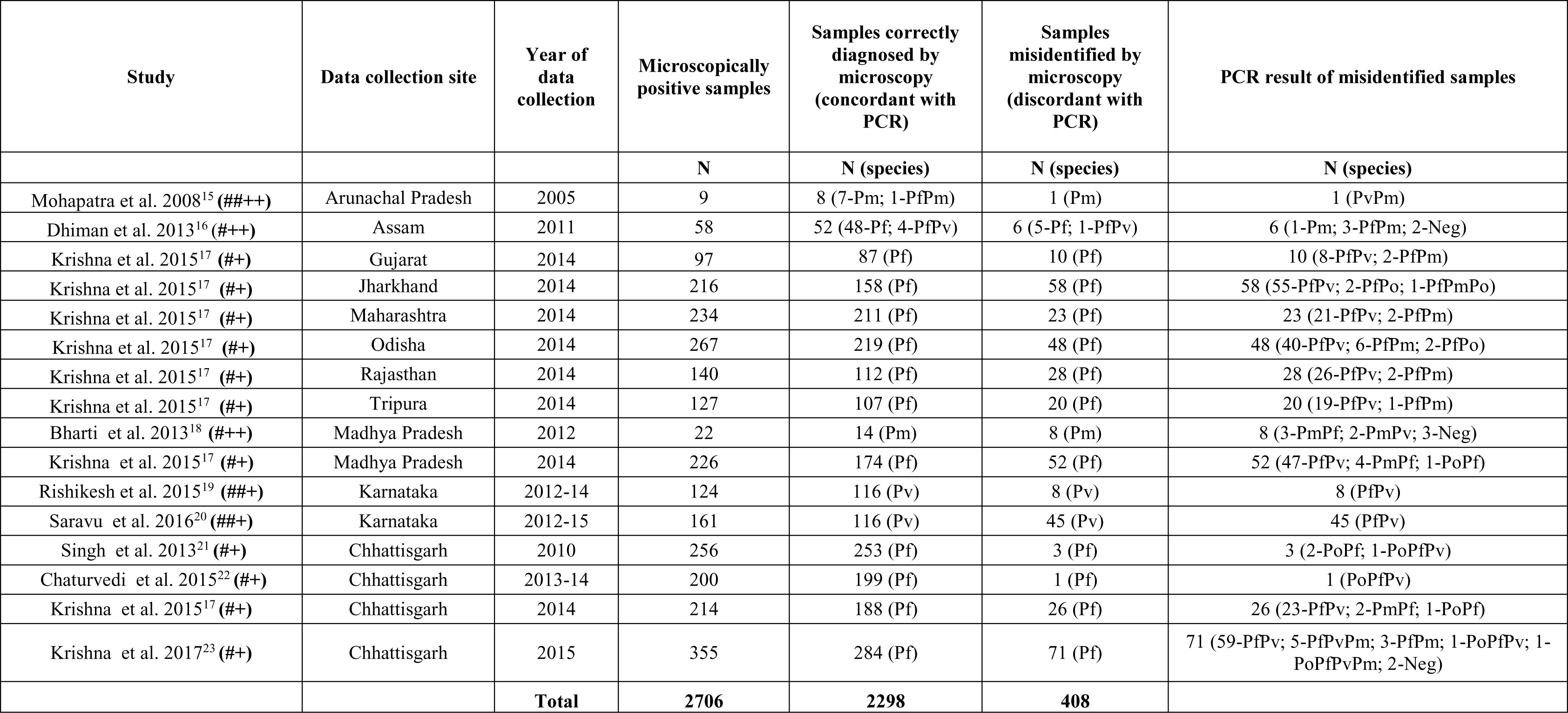
Detailed description of data analysed from all nine included reports. The table represents concordant and discordant results (species-wise) of microscopy and PCR. Additional information: Except for one study (Bharti et al. 2013), none of them had enough information to label their microscopists as “Experts”. Further, the type of study design was marked as (##) Longitudinal, (#) Cross-sectional, (++) Active & (+) Passive.

As expected, Pf and Pv were correctly identified by MS but only when present as mono-infections (Table 1 & Figure 2a) and not as mixed-infections with other *Plasmodium* species wherein one of the species was found missing by microscopy. As shown in Table 1, MS-false positives are consistent in Pf and Pv mono-infections across all studies, whereas MS-false negatives are restricted to almost all mixed-species infections, including PfPv. In was quite surprising that MS misidentified >98% (400/405) of mixed-infections as mono-infections, except PfPm and PfPv.^15–16^

**Figure 2a:**
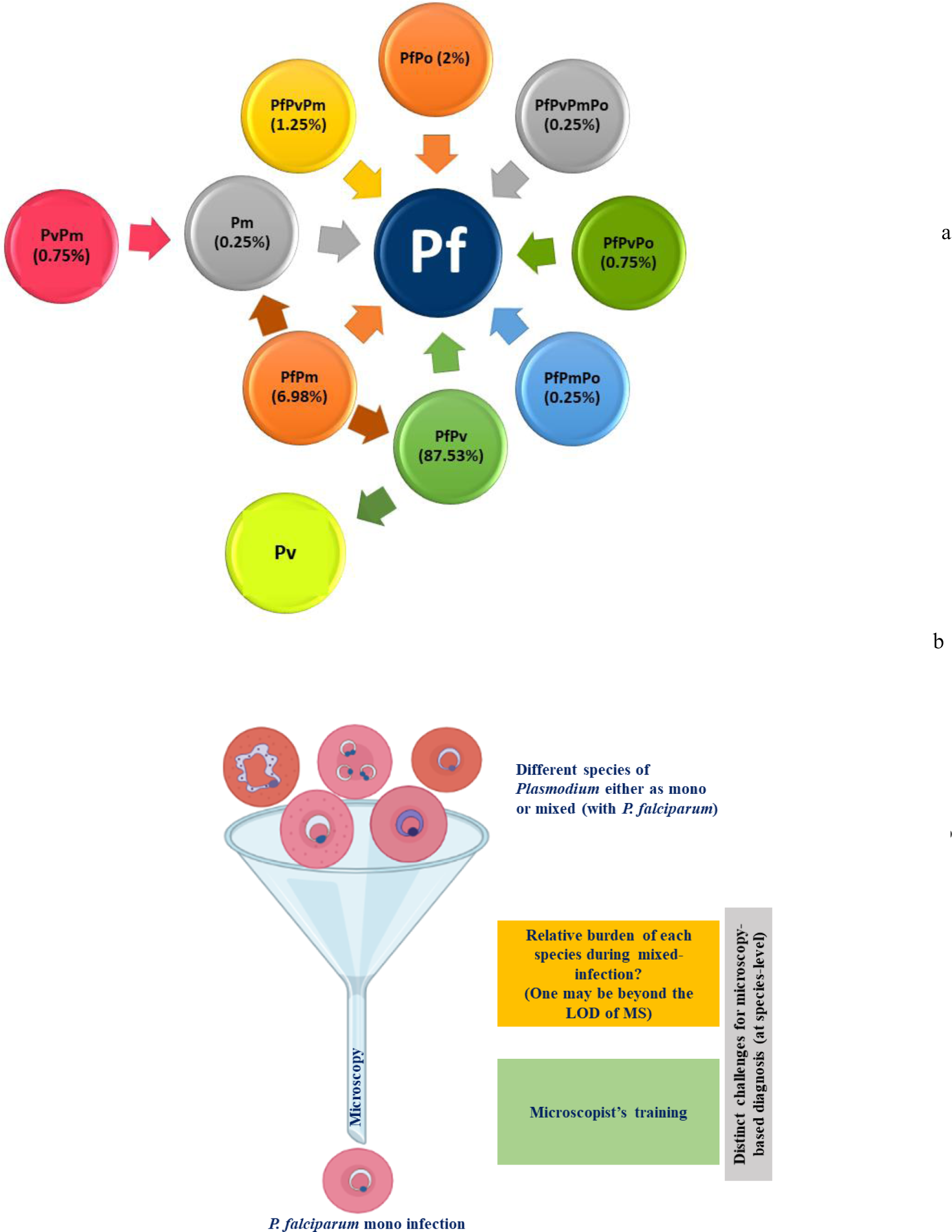
Microscopy v/s PCR outcome. The figure was made using data extracted from the enrolled studies & percentages were calculated as formula B2. The arrow heads represent microscopy outcome, whereas the arrow tails represent PCR outcome from different reports. Here, the percentages represent proportion of their misidentification to either mixed or some other species by microscopy. **Figure 2b: Possible challenges for microscopy at species-level diagnosis.** The figure represents the difficulties for microscopy in diagnosing a non-falciparum species when it co-exists with *P. falciparum*.

Figure 2a depicts that almost all mixed *Plasmodium* species infections were misidentified as Pf mono-infection by MS. The analysis strongly suggests that Pf is the most familiar species to microscopists. Out of total MI cases, ∼88% of mixed PfPv infections were misidentified as Pf or Pv mono-infections, indicating a significant failure of a microscopist. Further, PfPm was the second most overlooked mixed-infection after PfPv, with 7% (28/401) misidentification as Pf mono-infection. Other species found with Pf as mixed-infection were misidentified in ≤2% of cases.

As observed from Figure 2a, *Plasmodium* mixed-infections contribute more to misidentification by microscopy. Figure 3 illustrates the situation better by plotting species-specific MI cases out of total positive cases (by MS) of that species. MS did not misidentify any of the Pf (n=2040) or Pv (n=232) mono-infections whereas one Pm case (5%;1/22) was misidentified by MS. In case of mixed-infections, however, the picture is completely reversed, with a MI rate of >95%.

**Figure 3:**
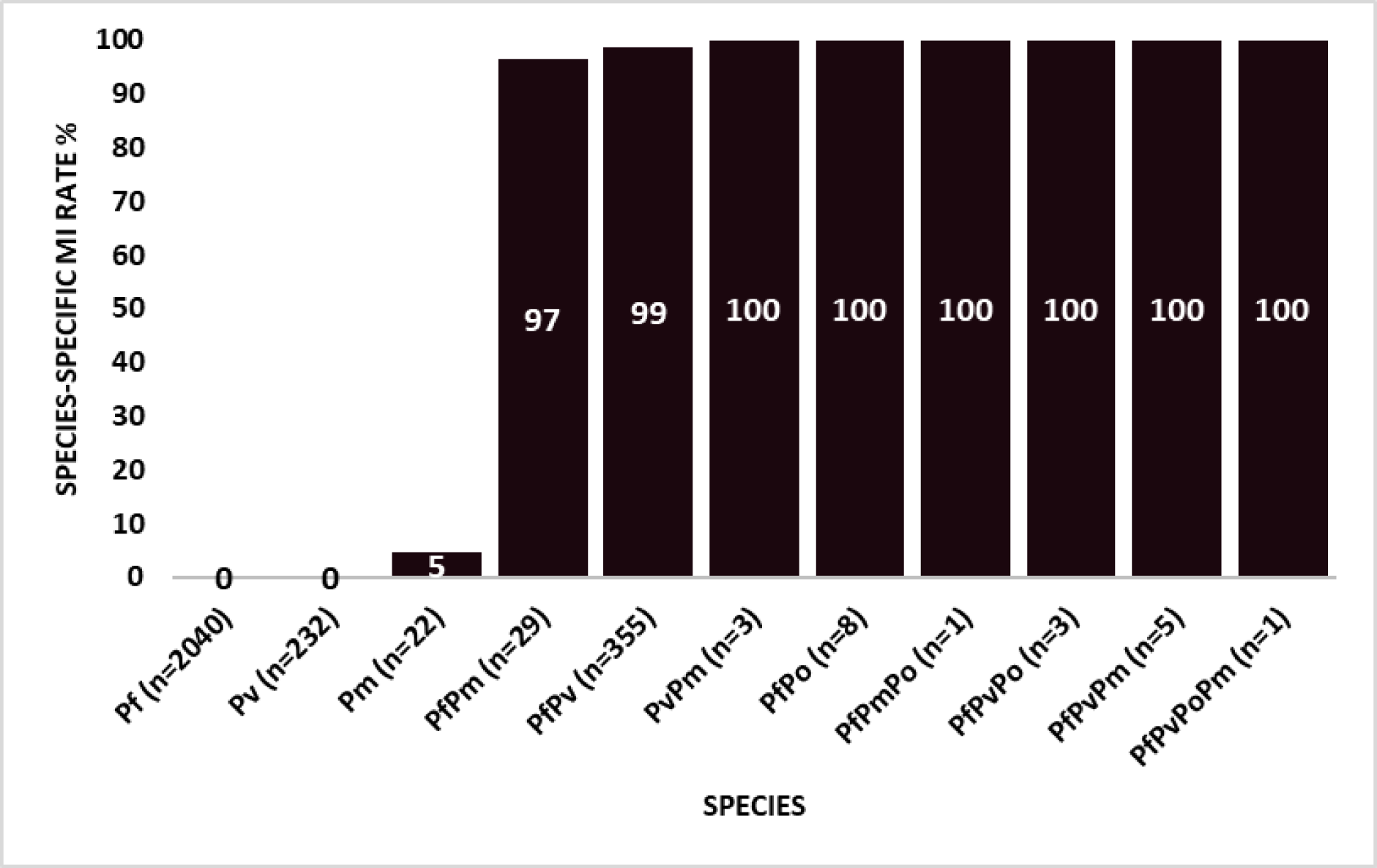
Contribution of *Plasmodium* species to microscopy misidentification. The figure demonstrates percentage of misidentification (by microscopy) of a particular species from confirmed positive cases (labelled as ‘n’) of that particular species detected by PCR (calculated as per formula B3).

Region-wise overall MI rates reveal that misidentification of *Plasmodium* species was highest in Jharkhand (27%; 58/216) followed by Madhya Pradesh (24%; 60/248), Rajasthan (20%; 28/140), Karnataka (19%; 53/285), Odisha (18%; 48/267) and Tripura (16%; 20/127). MI rate in the remaining five regions (Arunachal Pradesh, Assam, Gujarat, Chhattisgarh, and Maharashtra) ranged from 10 to 11% (Figure 4).

**Figure 4:**
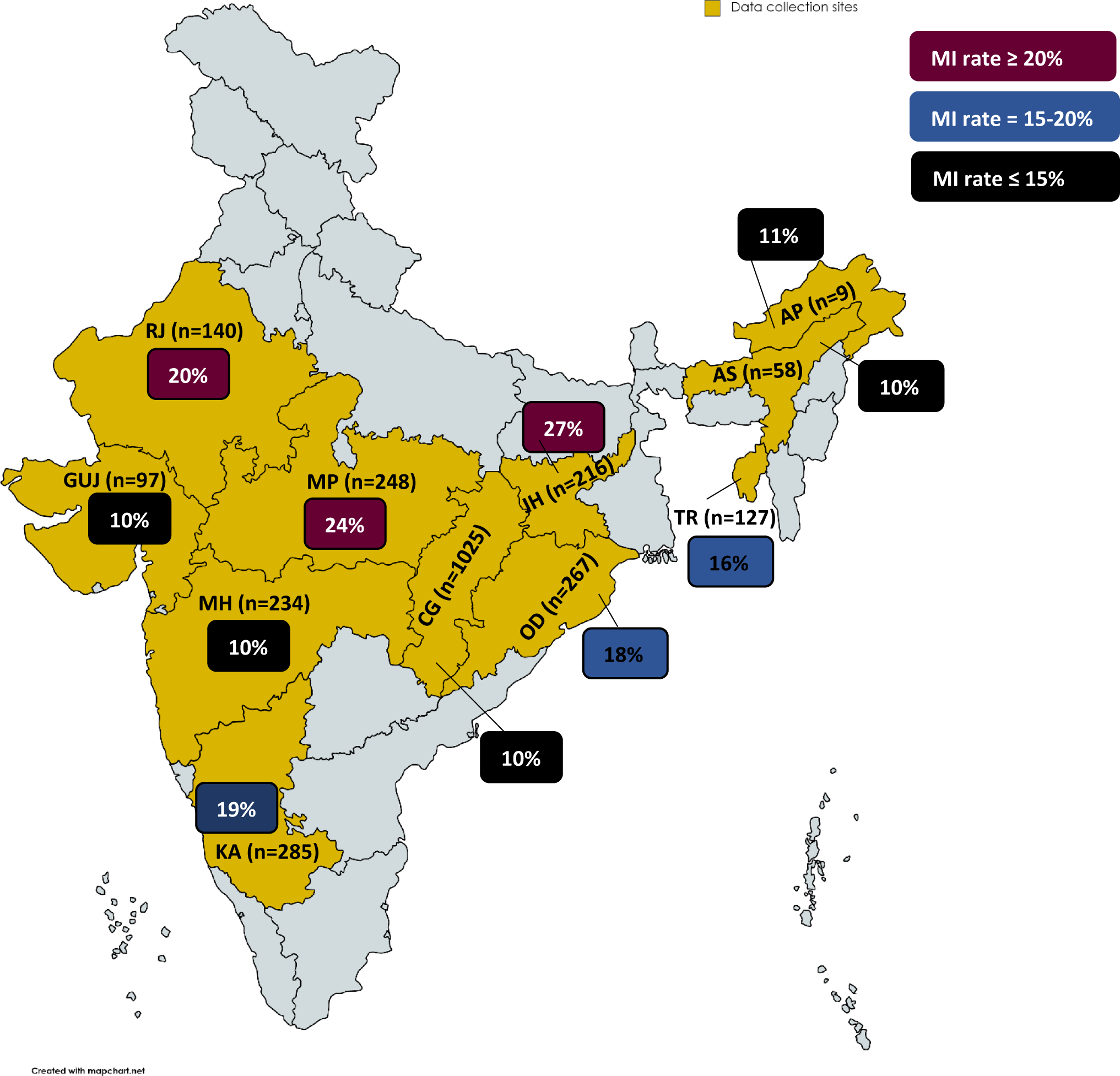
Geographical distribution of *Plasmodium* species misidentification (by microscopy). The figure depicts the contribution of various geographical sites (Indian states) to microscopy misidentification of *Plasmodium* species (calculated as formula A). Here, the yellow shaded areas indicate the geographic locations from which data have been gathered. These areas are labelled with the state name initials and the number (n) of samples that have tested positive for *Plasmodium* (by microscopy). According to their contribution, the percentages of MI (from the total positive samples for *Plasmodium*) from that specific region have been mentioned in boxes and coloured burgundy (MI rate 20%), blue (MI rate = 15-20%), and black (MI rate 15%). AP: Arunachal Pradesh, AS: Assam, CH: Chhattisgarh, GUJ: Gujarat, JH: Jharkhand, KA: Karnataka, MP: Madhya Pradesh, MH: Maharashtra, OD: Odisha, RJ: Rajasthan, TR: Tripura

## Discussion

Accurate diagnosis and radical cure is crucial in malaria elimination settings. This study results revealed that when a non-Pf species coexists with Pf as mixed-infection, it is misidentified as Pf mono-infection by MS in >95% of the cases, indicating a significant failure in reporting species-specific prevalence (Figures 2a & 3, 4). Although, it is possible that the other misidentified species present with Pf in mixed-infection were below the MS LoD, this cannot be true to all MI cases. In addition, it is difficult to accept that issues related to the sensitivity of MS are always associated with non-falciparum species. Similar findings were observed during a malaria MS remedial course conducted for ten days in Kenya and Ghana wherein MI was frequent with non-falciparum species.^25^ MS has its own challenges at species-level diagnosis both in terms of the technology (threshold; LoD) and the miscroscopist (training) which are represented in Figure 2b.

Another possibility that could exist is the morphological similarities between two different *Plasmodium* species making misidentification quite possible. Late stages trophozoites, schizonts, and gametocytes of *P. knowlesi* and *P. malariae* appear similar on blood smear, making differentiation difficult. Young rings of *P. knowlesi* also resemble with *P. falciparum* having double chromatin dots.^26^ Both *P. ovale* and *P. vivax* infect immature erythrocytes, and their infected cells have amoeboid trophozoites with schüffner’s dots. A multi-country meta-analysis was done to rule out Po misidentified as Pv & found that 11% of Po cases were misidentified as Pv in routine diagnosis.^7^ Another multi-country systematic review conducted to quantify the MI rate of Pk as Pm & reported 57% pooled prevalence.^8^ Such outcomes indicate the need of molecular assays to unveil the real species-specific load and to identify whether the species-specific parasite load is eligible to be classified as a sub-microscopic infection or not.

Evidence exists that raise such issues in piecemeal indicating poor MS-based diagnosis and highlighting the need for trained and experienced microscopists in field settings.^27–28^

In an elimination setting, knowing each and every *Plasmodium-*infected individual is of paramount significance because it may lead to further parasite transmission and sustenance of an infectious reservoir in the absence of apparent disease burden that bears the potential of generating clinical cases, putting successful elimination at constant risk. ^29–31^ The major limitations with MS are its sensitivity and specificity which are driven by the skills of the microscopist and the technology *per se*. The differential skills of microscopists impart “subjectivity” in malaria diagnosis which becomes a critical deterrent parameter. This subjectivity not only reflects in generating a range of sensitivities in detection of *Plasmodium* in blood smears but also compromises the specificity of MS in terms of identifying the parasite species correctly leading to misidentification of parasite species. The combinatorial effect of a compromised sensitivity and specificity thus reflects in a high burden of sub-microscopic infections in a region. In the context of species-specific diagnosis, this might under-estimate the burden of certain “cryptic” parasite species which were not known to exist or exist as a rare species which has many downstream clinical and therapeutic ramifications.

### Challenges & way ahead

As an identified challenge, we would like to revisit the concept of false positive (FP) and false negative (FN) MS diagnosis because it is frequently misinterpreted at species level.

When a sample is investigated both by MS and nested PCR (nPCR) for *Plasmodium* genus and species, the discordant (FN & FP) MS results against nPCR demand special attention as these generate various possibilities that are most likely to be misinterpreted. We hereby suggest two further ramifications of FN results using an ultrasensitive qPCR (Figure 5). As evident from the figure, qPCR will tend to classify MS FN samples into SMIs and actual FN where specific interventions could be targeted. Similarly, MS FP results can be refined by using a pan-*Plasmodium* nPCR into actual FP and species misidentification where both the scenarios would demand remedial training of micrscopist. It is also pertinent to say here that such granularity and factuality may be deciphered when each MS and PCR sample pair is analysed one-to-one.

**Figure 5:**
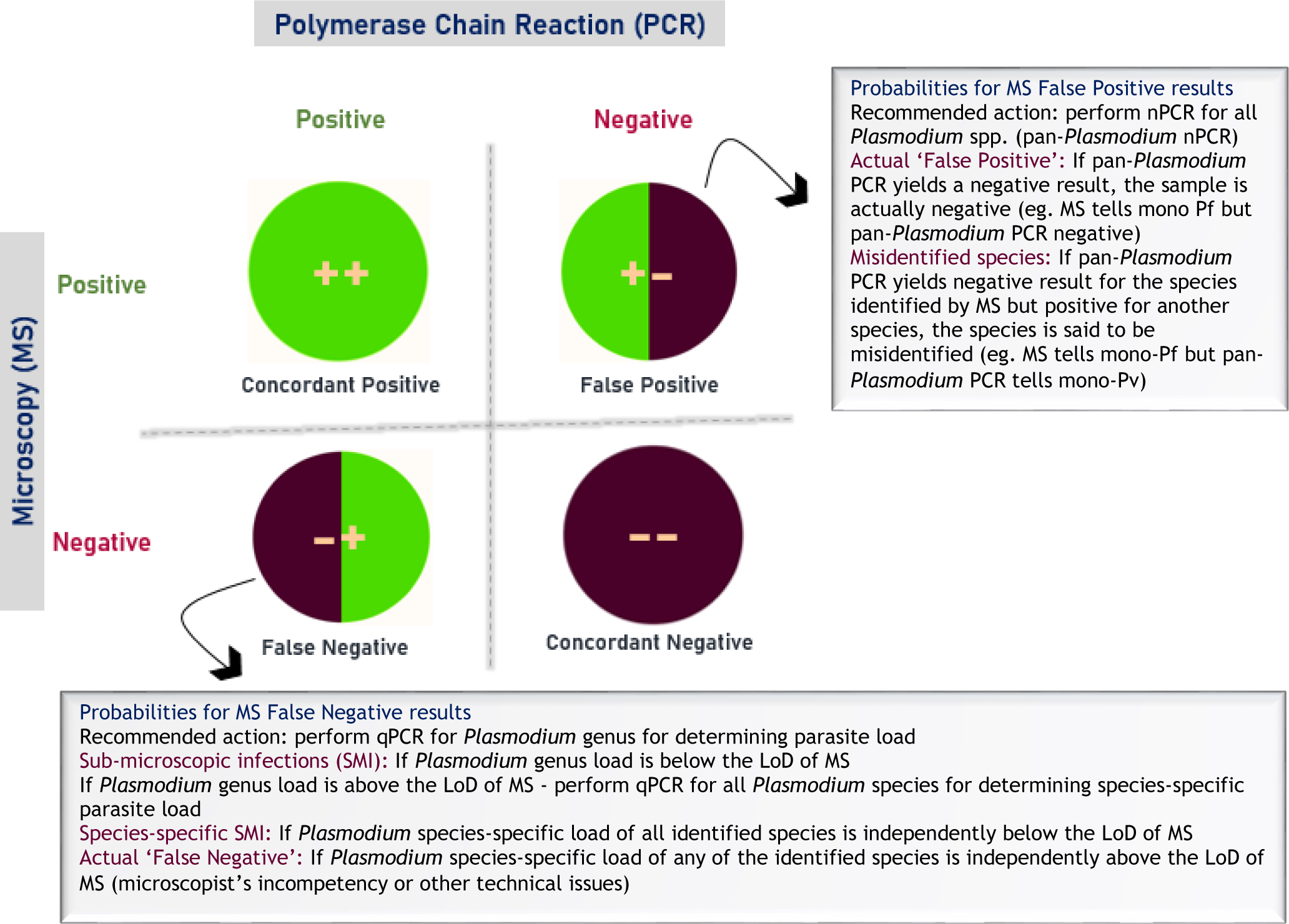
Four possibilities of Microscopy and PCR paired results for the same sample for *Plasmodium* parasite genus and species. The figure reframes the two common probabilities of a false microscopy and PCR pair results with a new and detailed perspective. It also suggests recommendations for obtaining more factual conclusions from the false positive and false negative outcomes that might otherwise be frequently misinterpreted.

MS FN results, commonly called as SMIs, can be estimated if the combinatorial result of MS and PCR is available for a particular sample whereas SMIs are commonly estimated from the paired diagnoses of all samples taken together (overall analysis) and not by independent sample paired analysis (one-to-one analysis). This is a real challenge as it tends to impart errors in actionable information interpreted from such data analysis as exemplified in Figure 6. The figure puts forth three hypothetical scenarios examining 5 independent samples in each by MS and PCR, independently.

**Figure 6:**
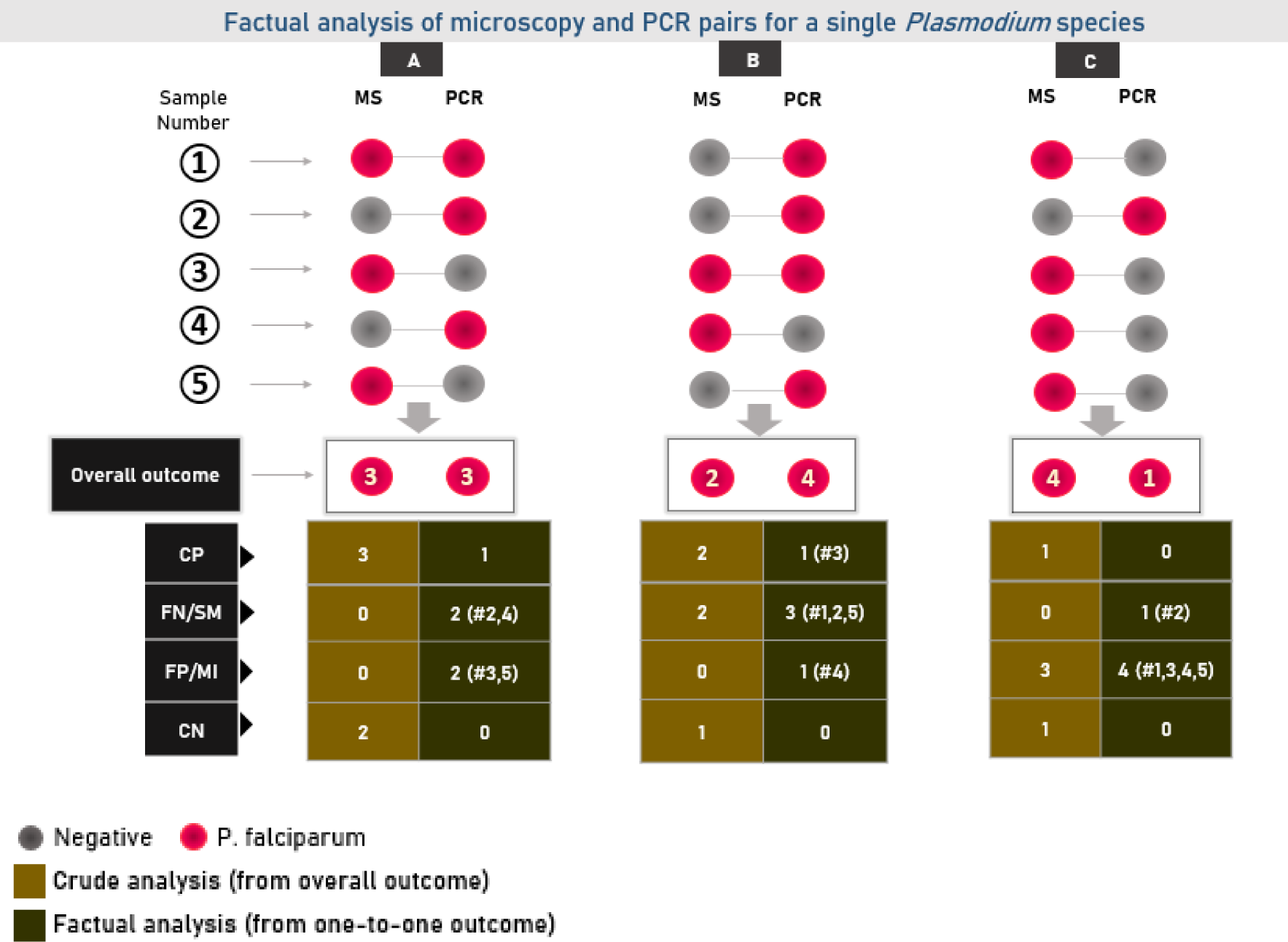
Differential interpretations from crude (overall outcome) and factual (one-to-one outcome) analyses of microscopy and PCR paired samples results revealing various pitfalls in identification of false negative (by microscopy) or sub-microscopic infections. The figure depicts how interpreting the number of false negative or sub-microscopic infections from an overall analysis vs one-to-one analysis might lead to erroneous interpretation of results. Presented here are 3 possible and different scenarios (A-C) wherein the microscopy (MS) and PCR results of 5 independent samples (1-5) are mentioned. The pink and grey circles denote positive and negative results by the two methods, respectively. For the ease of interpretation, a positive result by both microscopy (MS) and PCR is assumed to be of the same *Plasmodium* species, i.e. *P. falciparum*. CP (concordant positive) means “identical” microscopy and PCR results, whereas FN (false negative) and FP (false positive) are used to represent sub-microscopic infections and misidentified species, respectively. Concordant negative samples are demonstrated as CN for both microscopy and PCR. The comparative detailed interpretation from overall and one-to-one analyses is also described.

From an overall analysis of MS and PCR results from scenario A (total number of samples positive by each diagnostic method), it appears that the MS and PCR results are in perfect concordance (3 samples positive each by MS and PCR) with no SMI. However, in contrast, one-to-one pairwise sample analysis reveals the factual information that only 1 sample (sample #1) had concordance, two samples (samples #2 and 4) were SMI and two were FP by MS (samples #3 and 5). Similarly, the overall analysis from scenarios B and C reflects 2 SMIs and 3 MS FP diagnoses, respectively. The pairwise analysis reveals much striking difference in scenario B (1 concordant; 3 SMI and 1 FP) and C (1 SMI and 4 FP). The interpretation becomes very critical while taking a corrective action as no corrective action is apparently needed for scenario A whereas wrong corrective measures will be adopted from scenarios B and C, if the overall analysis was followed. This becomes much more important as the remedies depend on the type of error – SMIs might be addressed by rectifying issues related to microscopy (missing the parasite) unless the parasite load is below the LoD of MS and a FP diagnosis might have roots in misidentifying a *Plasmodium* species or detecting an artefact as a parasite. Since most published literatures present their results as “overall analysis”, it becomes difficult to get the real picture. Results from pairwise analysis of independent samples are thus recommended while reporting the outcomes from MS and PCR performed on the same sample.

The issues discussed above become even more complex when species-specific SMIs are investigated (Figure 7). At species level, mixed *Plasmodium* species infections provide distinct challenges for MS-based diagnosis for a variety of reasons. If two species coexist inside the same host, the relative burden of each species remains unknown; one may be beyond the LoD of MS while the other may not (SMI). In such situations, mixed-species infections are usually misdiagnosed as mono-species infections, resulting in a misleading diagnosis and treatment. On the other hand, if the burden of both the species is beyond MS LoD, it may be possible that the microscopists quit further reading of the slide as soon as s/he notices one of the two species. In the absence of independent sample pairwise data from published reports (as suggested here), the true picture remains under cover. The species-specific SMI data is critical for malaria control programs not only to identify the precise training needs but also to know if any particular *Plasmodium* species diagnosis is getting neglected thus leading to a building up of a reservoir with a tendency to emerge as a sudden outbreak.

**Figure 7:**
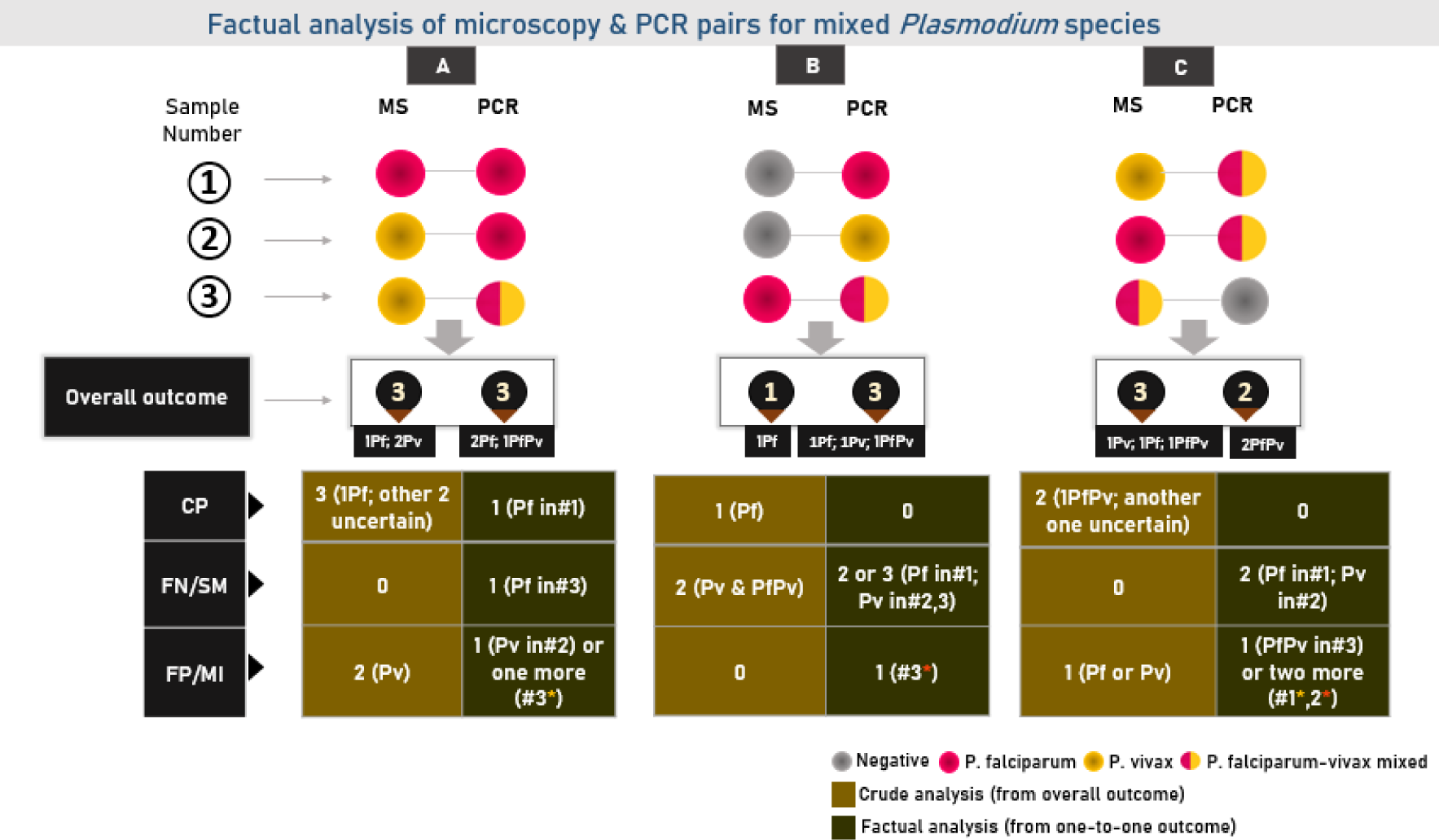
Depiction of the complexity associated with mixed *Plasmodium* species in interpreting false negative (by microscopy) or sub-microscopic infections from an overall sample analysis versus one-to-one sample analysis. Three different scenarios (A-C) are presented here, each with the MS and PCR results of three independent samples (1-3). The pink, yellow, and grey circles represent the Pf, Pv, and negative results obtained by the two methods, respectively. Samples infected with both Pf and Pv are represented by circles that contain both pink and yellow. CP denotes concordant positive or “identical” microscopy and PCR results, whereas FN and FP denote sub-microscopic and misidentified infections, respectively. This figure does not include any concordant negative samples. Samples with yellow asterisk show a possibility that Pf may have been misidentified as Pv, although Pv was also detected by PCR in that particular sample as a co-infecting species with Pf. Similarly, samples with orange asterisk demonstrate the opposite.

The database for the current study was restricted to *Plasmodium* mixed-infection, but the authors recommend that more studies should be conducted on *Plasmodium* mono-infections misidentification (by microscopy) to obtain a more complete picture. Because, in the case of mixed-infections that are misidentified as mono-infections, one species may be below the MS detection threshold, but this can be avoided in the case of mono-infections. In addition, because the included reports lack information on the type of microscopists, the authors are unable to identify whether an expert microscopist was involved or not. It also lacks studies involving Pk because they were omitted from the database owing to insufficient details.It has been noted that non-falciparum species (particularly *P. knowlesi*, *P. ovale curtisi* and *P. ovale wallikeri*) have rarely been diagnosed using molecular tools, which is a requirement in the current scenario, particularly in elimination settings.

## Conclusive remarks

*Plasmodium* misidentification by MS, coupled with the problem of SMI, are recognized yet under-reported problems in malaria-troubled countries. Both these attributes are the results of FP and FN MS-based malaria diagnosis, respectively, and have the potential to building up of undetected species-specific *Plasmodium* reservoirs that have a transmission potential and therefore thwart malaria elimination. Majority of published literature that reports misidentification and SMIs includes research not targeted specifically for quantifying misidentification and SMIs and report the same as a secondary or even a tertiary outcome. Those that do mention misidentification and SMIs often tend to report them in a flawed manner by adopting the summative analysis of microscopy-PCR paired results as shown here. The proposed one-to-one factual analyses of the MS-PCR paired samples helps in deciphering the misidentification and SMI data in a more granular way generating targeted actionable information for the program managers.

This research suggest that we might be overestimating the burden of *P. falciparum*, potentially misdirecting scarce elimination resources, while at the same time, underestimating non-falciparum species.

## Data Availability

All data produced in the present work are contained in the manuscript. In addition, the methods used to create the database that was used to include studies for this manuscript can be found here https://doi.org/10.1016/j.lansea.2022.05.001.

## Acknowledgement

Department of Science and Technology, Government of India is acknowledged to support ND through Women Scientist-A Scheme. ICMR-NIMR is acknowledged for infrastructural and administrative support

## Funding

Not applicable

## Conflict of interest

The authors declare no conflict of interest for the current study

## Authors’ contributions

Nimita Deora: Literature search, data acquisition, analysis and interpretation, drafting the manuscript and editing the manuscript critically, and figures

Dr Veena Pande: Supervision, analysis and interpretation, review & editing the manuscript critically

Dr Abhinav Sinha: Conceptualization, supervision, analysis and interpretation, review & editing the manuscript critically

## Notes

### Competing Interest Statement

The authors have declared no competing interest.

### Funding Statement

This study did not receive any funding

### Author Declarations

The study is a secondary data analysis of published studies in the field

